# Investigation of association of rare, functional genetic variants with heavy drinking and problem drinking in exome sequenced UK Biobank participants

**DOI:** 10.1101/2021.02.04.21251145

**Authors:** David Curtis

**Affiliations:** UCL Genetics Institute, UCL, Darwin Building, Gower Street, London WC1E 6BT; Centre for Psychiatry, Queen Mary University of London, Charterhouse Square, London EC1M 6BQ

**Keywords:** Alcohol, exome, *FOXP1*, *ARHGAP33*, *CDH9*, *VGF*

## Abstract

**Aims:** The study aimed to identify specific genes and functional genetic variants affecting susceptibility to two alcohol related phenotypes: heavy drinking and problem drinking.

**Methods:** Phenotypic and exome sequence data was downloaded from the UK Biobank. Reported drinks in the last 24 hours was used to define heavy drinking while responses to a mental health questionnaire defined problem drinking. Gene-wise weighted burden analysis was applied, with genetic variants which were rarer and/or had a more severe functional effect being weighted more highly. Additionally, previously reported variants of interest were analysed inidividually.

**Results:** Of exome sequenced subjects, for heavy drinking there were 8,166 cases and 84,461 controls while for problem drinking there were 7,811 cases and 59,606 controls. No gene was formally significant after correction for multiple testing but three genes possibly related to autism were significant at p < 0.001, *FOXP1, ARHGAP33* and *CDH9*, along with *VGF* which may also be of psychiatric interest. Well established associations with rs1229984 in *ADH1B* and rs671 in *ALDH2* were confirmed but previously reported variants in *ALDH1B1* and *GRM3* were not associated with either phenotype.

**Conclusions:** This large study fails to conclusively implicate any novel genes or variants. It is possible that more definitive results will be obtained when sequence data for the remaining UK Biobank participants becomes available and/or if data can be obtained for a more extreme phenotype such as alcohol dependence disorder. This research has been conducted using the UK Biobank Resource.

**Short summary:** Tests for association of rare, functional genetic variants with heavy drinking and problem drinking confirm the known effects of variants in *ADH1B* and *ALDH2* but fail to implicate novel variants or genes. Results for three genes potentially related to autism suggest they might exert a protective effect.

## Introduction

There is a heritable contribution to the related phenotypes of heavy drinking, problematic drinking and alcohol dependency disorder and this has been explored in a number of ways. As reviewed recently, genome wide association studies (GWAS) using common variants produce signals at a number of genes including some involved in alcohol metabolism, *ADH1B, ADH1C* and *AD4*, along with others with less obvious roles, *DRD2, KLB, CADM2, CRHR1, FGF14, GCKR, SIX3, SLC39A8* and *FTO* (Johnson et al., 2020). A subsequent GWAS of heavy alcohol consumption replicated some of these and additionally implicated the locus for a pseudogene, *BTF3P13* (Thompson et al., 2020).

GWAS results typically implicate genomic regions where common variation is associated with moderate differences in susceptibility to a trait but it is often difficult to elucidate which individual genes and variants are responsible and the biological mechanisms involved. In this respect, alcohol intake is somewhat exceptional among complex traits in that there are individual functional variants which have been well characterised. These are in genes involved in the metabolism of alcohol and consist of a non-synonymous gain of function variant in *ADH1B*, rs1229984, and a damaging variant in *ALDH2*, rs671, which both reduce the risk development of alcohol-related disorders by increasing the levels of circulating acetaldehyde following alcohol ingestion (Edenberg and McClintick, 2018). As they discuss, other variants in *ADH* genes have also been reported to be associated in some studies but linkage disequilibrium (LD) relationships in this region make interpretation difficult and likewise it is not clear if other variants in aldehyde dehydrogenase genes, including rs2228093 and rs2073478 in *ALDH1B1*, have effects. We have reported that a Kozak sequence variant in *GRM3*, rs148754219, is associated with alcohol dependence as well as schizophrenia and bipolar disorder but this finding has to date not been replicated (O’Brien et al., 2014).

The UK Biobank consists of a sample of 500,000 participants who have provided DNA samples and who have undergone phenotyping on a wide variety of measures on which GWAS have been performed, including the study of heavy drinking referred to above (Thompson et al., 2020). Exome sequence data has now been made available for 200,000 participants (Szustakowski et al., 2020). This makes it possible to investigate whether there may be rare sequence variants which have major effects on gene function and whether such variants within a gene are collectively associated with a trait of interest. Potentially the identification of rare variants with functional effects could facilitate biological insights. Such analyses successfully implicated functional genes in the case of hyperlipidaemia and type 2 diabetes but did not produce any positive results for a phenotype related to common mental illness (Curtis, 2021a, 2021b, 2021c).

Here, we use this approach to carry out weighted burden analysis of rare, functional variants on two alcohol-related phenotypes, heavy drinking and problem drinking.

## Methods

The UK Biobank dataset was downloaded along with the variant call files for 200,632 subjects who had undergone exome-sequencing and genotyping by the UK Biobank Exome Sequencing Consortium using the GRCh38 assembly with coverage 20X at 95.6% of sites on average (Szustakowski et al., 2020). UK Biobank had obtained ethics approval from the North West Multi-centre Research Ethics Committee which covers the UK (approval number: 11/NW/0382) and had obtained informed consent from all participants. The UK Biobank approved an application for use of the data (ID 51119) and ethics approval for the analyses was obtained from the UCL Research Ethics Committee (11527/001). All variants were annotated using the standard software packages VEP, PolyPhen and SIFT (Adzhubei et al., 2013; Kumar et al., 2009; McLaren et al., 2016). To obtain population principal components reflecting ancestry, version 2.0 of *plink* (https://www.cog-genomics.org/plink/2.0/) was run with the options --*maf 0.1 --pca 20 approx* (Chang et al., 2015; Galinsky et al., 2016).

The UK Biobank sample contains 503,317 subjects of whom 94.6% are of white ethnicity. As we have discussed previously, it has become standard practice for investigators to simply discard data from participants with other ancestries and we regard this as regrettable (Curtis, 2021d). We demonstrated that if population principal components are included as covariates then it is possible to include all participants, regardless of ancestry, in the type of weighted burden analysis described here without any inflation of the test statistic.

Two alcohol related phenotypes were assigned as follows. The phenotype for heavy drinking was defined in a similar way to that used in the previous GWAS using 125,249 UK Biobank participants, except that was based on reported average drinks in a week whereas instead we used reported drinks in the previous 24 hours (Thompson et al., 2020). On a number of occasions a subset of participants were asked about their dietary intake in the previous 24 hours and if they had consumed alcohol they were asked to provide the number of drinks for different types of beverage. From these reports, the total daily intake was then estimated using the same number of units per drink as in the previous study: beer/cider = 2.6; white wine = 1.5; red wine = 1.5; fortified wine = 1.1; spirits = 1; other = 1.5. If 7 times this daily intake exceeded 50 for males or 35 for females the participant was defined as a case in respect of heavy drinking, whereas controls were taken as participants who had completed the questionnaire on at least one occasion but did not exceed these limits.

The phenotype of problem drinking was based on the 157,379 participants who completed a mental health questionnaire. Cases were defined as participants answering Yes to any of these three questions:

> Have you been physically dependent on alcohol?
>
> Has a relative or friend or a doctor or another health worker been concerned about your drinking or suggested you cut down?
>
> Have you been addicted to alcohol?

Participants selecting a response of Monthly or more frequently to any of these questions were also classified as cases:

> How often during the last year have you failed to do what was normally expected from you because of drinking?
>
> How often during the last year have you been unable to remember what happened the night before because you had been drinking?
>
> How often during the last year have you had a feeling of guilt or remorse after drinking?
>
> How often during the last year have you needed a first drink in the morning to get yourself going after a heavy drinking session?
>
> How often during the last year have you found that you were not able to stop drinking once you had started?

All other participants who had completed the mental health questionnaire were counted as controls.

The SCOREASSOC program was used to carry out a weighted burden analysis to test whether, in each gene, sequence variants which were rarer and/or predicted to have more severe functional effects occurred more commonly in cases than controls. Attention was restricted to rare variants with minor allele frequency (MAF) <= 0.01 in both cases and controls. As previously described, variants were weighted by overall MAF so that variants with MAF=0.01 were given a weight of 1 while very rare variants with MAF close to zero were given a weight of 10 (Curtis, 2021d). Variants were also weighted according to their functional annotation using the GENEVARASSOC program, which was used to generate input files for weighted burden analysis by SCOREASSOC (Curtis, 2016, 2012). The weights were informed from the analysis of the effects of different categories of variant in *LDLR* on hyperlipidaemia risk (Curtis, 2021a). Variants predicted to cause complete loss of function (LOF) of the gene were assigned a weight of 100. Nonsynonymous variants were assigned a weight of 5 but if PolyPhen annotated them as possibly or probably damaging then 5 or 10 was added to this and if SIFT annotated them as deleterious then 20 was added. In order to allow exploration of the effects of different types of variant on risk the variants were also grouped into broader categories to be used in multivariate analyses as described below. The full set of weights and categories is displayed in Table 1.

**Table 1.** The table shows the weight which was assigned to each type of variant as annotated by VEP, Polyphen and SIFT as well as the broad categories which were used for multivariate analyses of variant effects (Adzhubei et al., 2013; Kumar et al., 2009; McLaren et al., 2016).

| VEP / SIFT / Polyphen annotation | Weight | Category |
| --- | --- | --- |
| intergenic_variant | 0 | Unused |
| feature_truncation | 0 | Intronic, etc. |
| regulatory_region_variant | 0 | Intronic, etc. |
| feature_elongation | 0 | Intronic, etc. |
| regulatory_region_amplification | 1 | Intronic, etc. |
| regulatory_region_ablation | 1 | Intronic, etc. |
| TF_binding_site_variant | 1 | Intronic, etc. |
| TFBS_amplification | 1 | Intronic, etc. |
| TFBS_ablation | 1 | Intronic, etc. |
| downstream_gene_variant | 0 | Intronic, etc. |
| upstream_gene_variant | 0 | Intronic, etc. |
| non_coding_transcript_variant | 0 | Intronic, etc. |
| NMD_transcript_variant | 0 | Intronic, etc. |
| intron_variant | 0 | Intronic, etc. |
| non_coding_transcript_exon_variant | 0 | Intronic, etc. |
| 3_prime_UTR_variant | 1 | 3 prime UTR |
| 5_prime_UTR_variant | 1 | 5 prime UTR |
| mature_miRNA_variant | 5 | Unused |
| coding_sequence_variant | 0 | Unused |
| synonymous_variant | 0 | Synonymous |
| stop_retained_variant | 5 | Unused |
| incomplete_terminal_codon_variant | 5 | Unused |
| splice_region_variant | 1 | Splice region |
| protein_altering_variant | 5 | Protein altering |
| missense_variant | 5 | Protein altering |
| inframe_deletion | 10 | InDel, etc |
| inframe_insertion | 10 | InDel, etc |
| transcript_amplification | 10 | InDel, etc |
| start_lost | 10 | Unused |
| stop_lost | 10 | Unused |
| frameshift_variant | 100 | Disruptive |
| stop_gained | 100 | Disruptive |
| splice_donor_variant | 100 | Splice site variant |
| splice_acceptor_variant | 100 | Splice site variant |
| transcript_ablation | 100 | Disruptive |
| SIFT deleterious | 20 | Deleterious |
| PolyPhen possibly damaging | 5 | Possibly damaging |
| PolyPhen probably damaging | 10 | Probably damaging |

As described previously, the weight due to MAF and the weight due to functional annotation were multiplied together to provide an overall weight for each variant. Variants were excluded if there were more than 10% of genotypes missing in the controls or if the heterozygote count was smaller than both homozygote counts in the controls. If a subject was not genotyped for a variant then they were assigned the subject-wise average score for that variant. For each subject a gene-wise weighted burden score was derived as the sum of the variant-wise weights, each multiplied by the number of alleles of the variant which the given subject possessed. For variants on the X chromosome, hemizygous males were treated as homozygotes.

For each gene, logistic regression analysis was carried out including the first 20 population principal components and sex as covariates and a likelihood ratio test was performed comparing the likelihoods of the models with and without the gene-wise burden score. The statistical significance was summarised as a signed log p value (SLP), which is the log base 10 of the p value given a positive sign if the score is higher in cases and negative if it is higher in controls.

Gene set analyses were carried out as before using the 1454 “all GO gene sets, gene symbols” pathways as listed in the file *c5*.*all*.*v5*.*0*.*symbols*.*gmt* downloaded from the Molecular Signatures Database at http://www.broadinstitute.org/gsea/msigdb/collections.jsp (Subramanian et al., 2005). For each set of genes, the natural logs of the gene-wise p values were summed according to Fisher’s method to produce a chi-squared statistic with degrees of freedom equal to twice the number of genes in the set. The p value associated with this chi-squared statistic was expressed as a minus log10 p (MLP) as a test of association of the set with the hyperlipidaemia phenotype.

Individual coding variants which had previously been reported to be associated with alcohol-related phenotypes were entered into logistic regression analyses with principal components and sex as covariates. The variants in the *ADH* gene cluster were analysed jointly in order to avoid spurious results due to LD relationships.

For selected genes, additional analyses were carried out to clarify the contribution of different categories of variant. As described previously, logistic regression analyses were performed on the counts of the separate categories of variant as listed in Table 1, again including principal components and sex as covariates, to estimate the effect size for each category (Curtis, 2021e). The odds ratios associated with each category were estimated along with their standard errors and the Wald statistic was used to obtain a p value, except for categories in which variants occurred fewer than 50 times in which case Fisher’s exact test was applied to the variant counts. The associated p value was converted to an SLP, again with the sign being positive if the mean count was higher in cases than controls. In the case of *ADH1C*, the individual variants referred to above within the *ADH* region were included as additional covariates.

## Results

Only a subset of the exome sequenced subjects had completed the relevant questionnaires. For the heavy drinking phenotype there were 8,166 cases and 84,461 controls. For the problem drinking phenotype there were 7,811 cases and 59,606 controls. There were 20,384 genes for which there were qualifying variants. Given this and the fact that two phenotypes were tested, the critical threshold for the absolute value of the SLP to declare a result as formally statistically significant is −log10(0.05/(20384*2)) = 5.91 and no gene achieved this for either phenotype. Figure 1A shows the QQ plot for the heavy drinking phenotype, which shows that the SLPs conform well with the null hypothesis expectation. If the genes with the 100 highest and lowest SLPs, which might be capturing a biological effect, are disregarded then the gradient for the positive SLPs is 0.99 with intercept at −0.038 and the negative SLPs have gradient 1.03 with intercept at −0.042. Figure 1B shows the QQ plot for problem drinking. With the 100 highest and lowest SLPs removed the positive SLPs have intercept −0.032 and gradient 0.99 while the negative SLPs have intercept −0.046 and gradient 1.04. Given that the null hypothesis expectation is that the gradient should be 1 with intercept at 0, these results suggest that the approach used is sound and that including the covariates means that there is little inflation of the test statistics due to population stratification or other artefacts.

**Figure 1.**
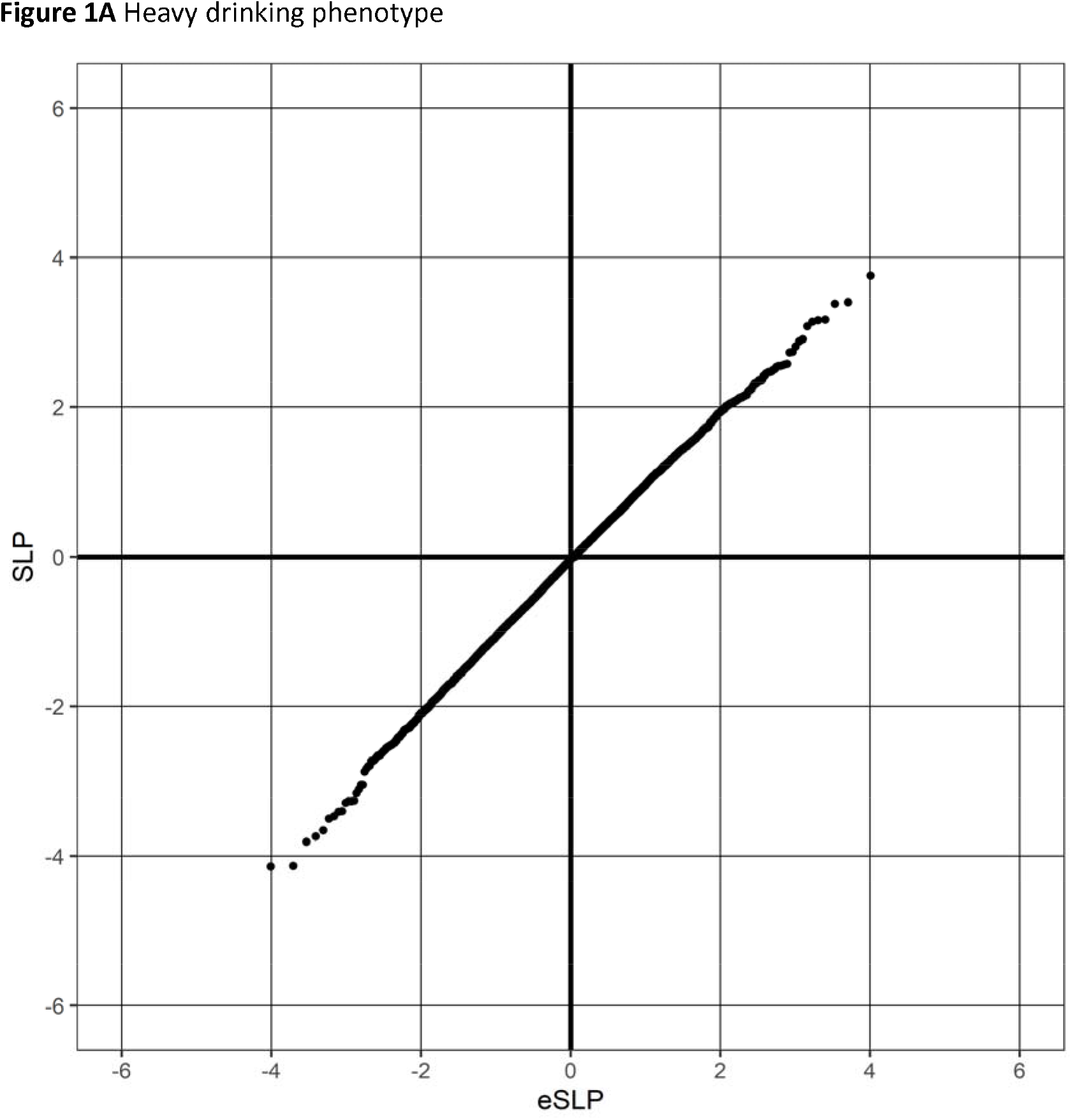
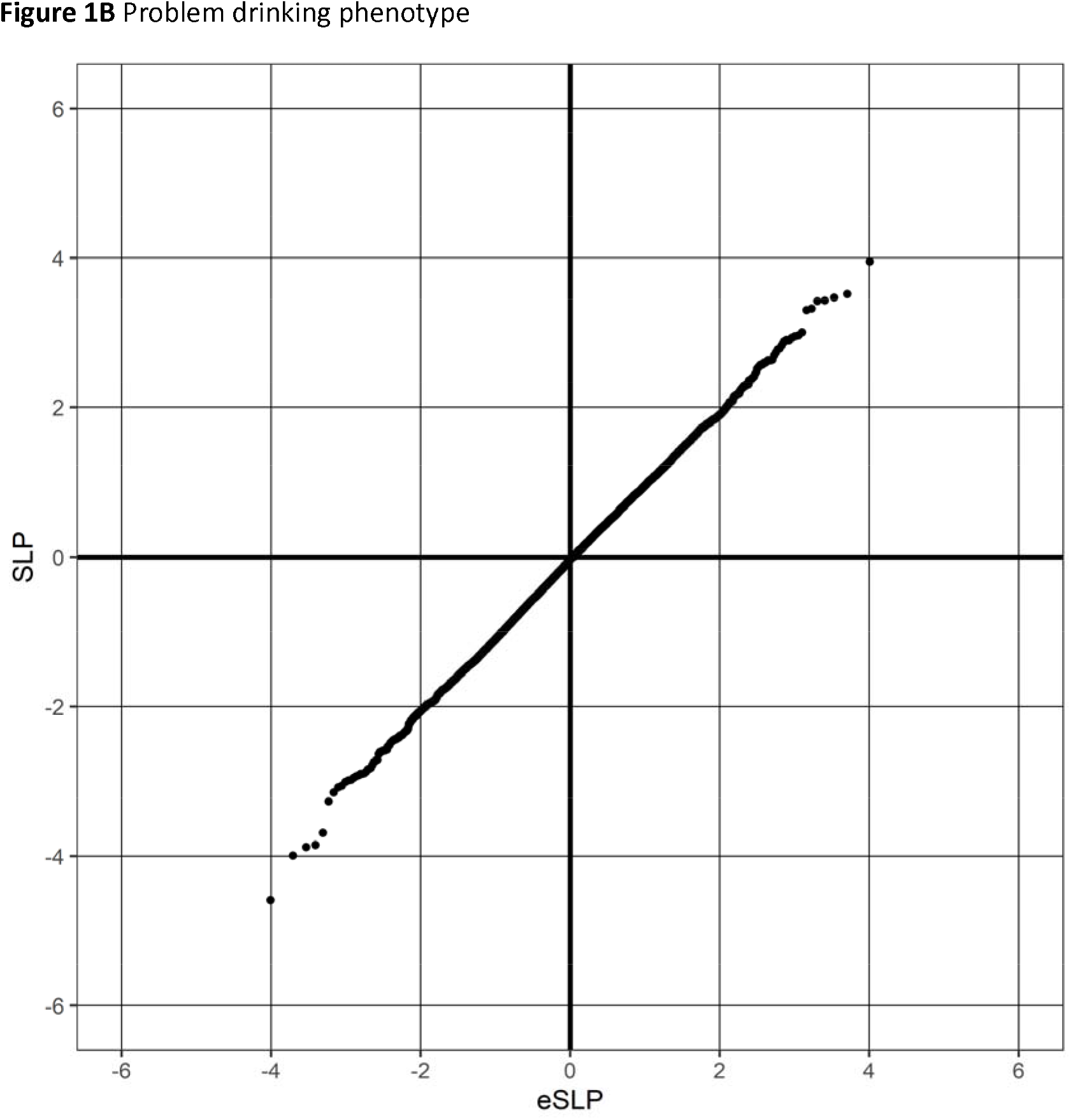
QQ plot of SLPs obtained for weighted burden analysis of association with alcohol phenotypes showing observed against expected SLP for each gene.

By chance, one would expect about 20 genes to have SLP with absolute value exceeding 3 (equivalent to p<0.001) for each phenotype whereas the actual numbers are 24 for heaving drinking and 18 for problem drinking. These genes are listed in Table 2 and, although not formally significant after correction for multiple testing, a few seem to be of potential interest.

**Table 2.**
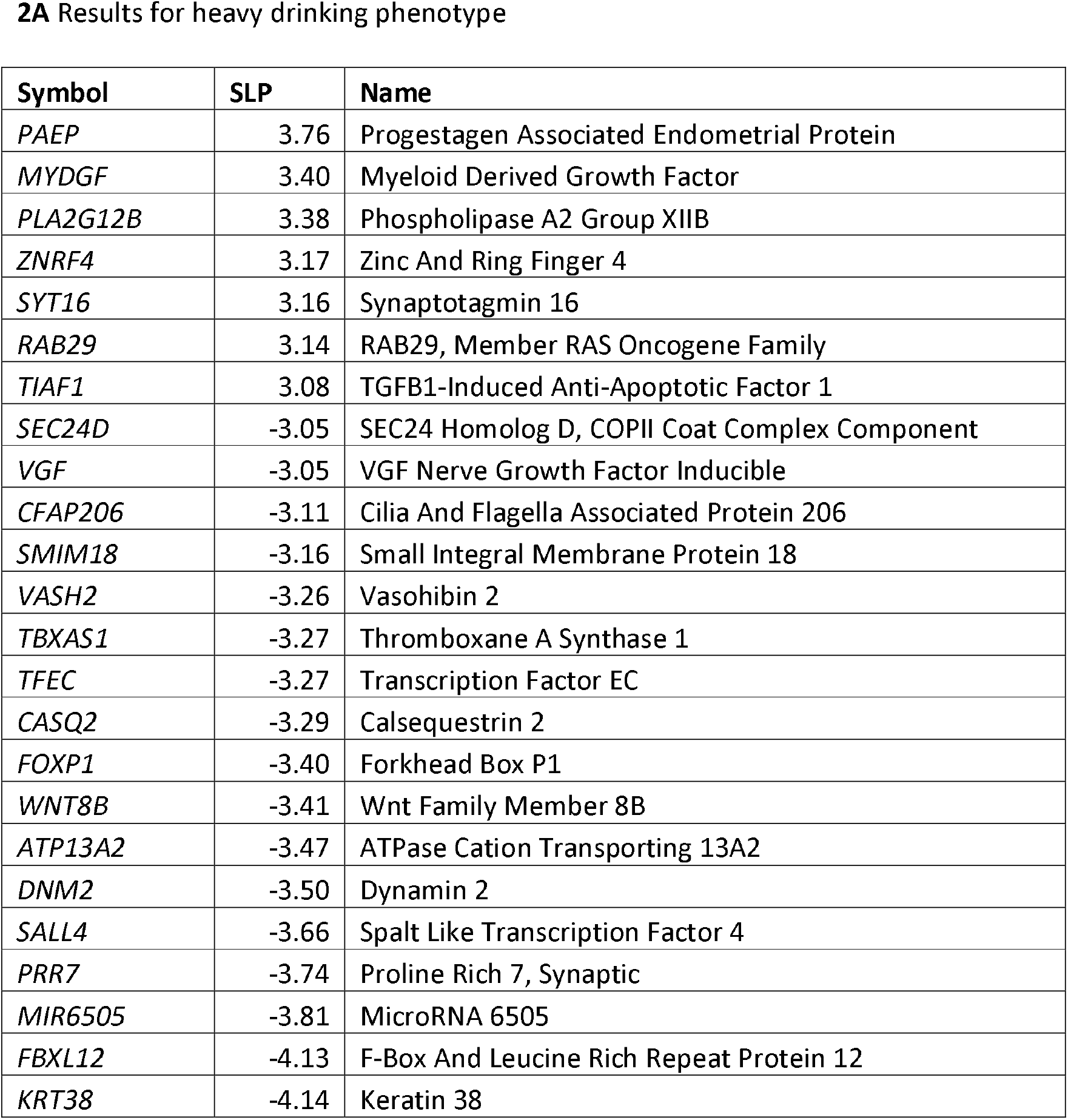
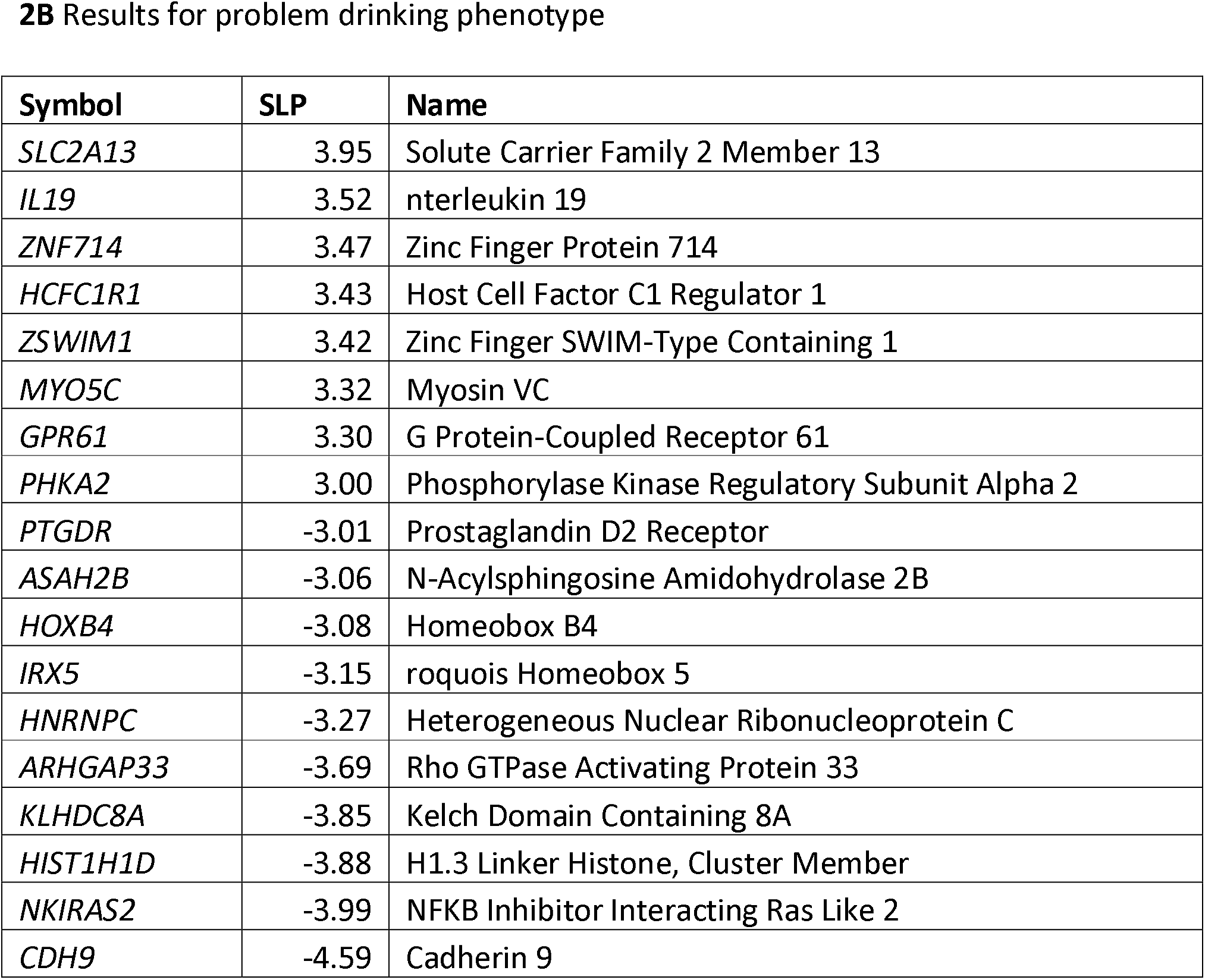
Genes with absolute value of SLP exceeding 3 or more (equivalent to p<0.001) for test of association of weighted burden score with alcohol phenotypes.

Using heavy drinking as the phenotype, *VGF* produces SLP = −3.05, suggesting that functional variants in it might be associated with lower alcohol intake. As reviewed recently, *VGF* is expressed extensively in central nervous system and there are reports that its expression is reduced in patients with depression but increased in schizophrenia, while both heterozygous *Vgf* knockout mice and *Vgf* over-expressing mice display some depressive features (Mizoguchi et al., 2019). *CFAP206* produces SLP = −3.11 but although expression of *Cfap206* is up-regulated in the hippocampus by alcohol administration in rats there seems little else to suggest that it is likely to affect alcohol intake (Choi et al., 2020). Disruption of *FOXP1* (SLP = −3.4) has been reported to be associated with cognitive dysfunction including intellectual disability, autism spectrum disorder and language impairment as well as psychiatric symptoms (Bacon and Rappold, 2012; Siper et al., 2017). The two genome wide significant hits in a study of self-reported childhood maltreatment in 124,711 UK Biobank participants were at *FOXP1* and *FOXP2*, leading the authors to suggest that these genes might be involved in externalising symptoms (Dalvie et al., 2020).

Two of genes with SLP < −3 for the problem drinking phenotype are also possibly implicated in autism risk, *ARHGAP33* (SLP = −3.69) and *CDH9* (SLP = −4.59). *ARHGAP33* is involved in dendrite and synapse development and its loss is associated with autism-like behaviours in mice (Nakazawa et al., 2016; Rosário et al., 2007; Schuster et al., 2015). A homozygous non-synonymous variant in *ARHGAP33* has been reported as a possible cause of a case of generalised developmental delay with seizures, microcephaly and dysmorphic features (Anazi et al., 2017). Common variants between *CDH9* and *CDH10* were shown to be associated with autism and it is now recognised that several cadherins are involved in neuronal development and are designated as autism risk genes (Lin et al., 2016; Redies et al., 2012; Wang et al., 2009).

In order to see if any additional genes were highlighted by analysing gene sets, gene set analysis was performed as described above. Given that 1,454 sets were tested for two phenotypes, a critical MLP to achieve to declare results significant after correction for multiple testing would be −log10(0.05/(1454*2)) = 4.76 and this was not achieved by any set. Two sets achieved MLP > 3 with the problem drinking phenotype, NEGATIVE REGULATION OF CYTOSKELETON ORGANIZATION AND BIOGENESIS (MLP = 4.27) and RESPONSE TO BIOTIC STIMULUS (MLP = 3.32). Inspection of the detailed results for these sets did not suggest any additional genes as plausible candidates.

None of the genes mentioned in introduction showed evidence for association with either phenotype using the gene-based burden tests, with the most significant result being SLP = 1.45 for GCKR with heavy drinking. The results for all genes are presented in Supplementary Table S1 and for all sets of genes in Supplementary Table S2.

The results for the variants previously reported to be associated with alcohol-related phenotypes are shown in Table 3. This shows that rs1229984, the gain of function variant in *ADH1B*, produces highly significant evidence for a protective effect for both phenotypes but that the other tested variants in *ADH1B* and *ADH1C* are not significantly associated with either phenotype. The damaging variant in *ALDH2*, rs671, is much less frequent in cases than controls for both phenotypes but is so rare in this largely European sample that it only produces modestly significant results. The tested variants in ALDH1B1 and *GRM3* have equal frequencies in cases and controls for both phenotypes.

**Table 3.** Results of analyses of individual variants previously reported to be implicated in alcohol related phenotypes, showing allele frequency in controls and cases. Population principal components and sex were included in all analyses in order to calculate the SLP. For the analyses of the variants in *ADH1B* and *ADH1C*, the SLP was derived from a multivariate analysis which included all variants in these genes simultaneously.

| Variant | Gene | Chr | Position | Heavy drinking |  |  | Problem drinking |  |  |
| --- | --- | --- | --- | --- | --- | --- | --- | --- | --- |
|  |  |  |  | Control allele frequency | Case allele frequency | SLP | Control allele frequency | Case allele frequency | SLP |
| rs1229984 | *ADH1B* | 4 | 99318162 | 0.0345210 | 0.018185 | -14.00 | 0.034420 | 0.017219 | -16.48 |
| rs2066702 | *ADH1B* | 4 | 99307860 | 0.0044820 | 0.002265 | -0.32 | 0.003003 | 0.001536 | -0.69 |
| rs698 | *ADH1C* | 4 | 99339632 | 0.4008950 | 0.418677 | -0.19 | 0.403057 | 0.420305 | 0.16 |
| rs1693482 | *ADH1C* | 4 | 99342808 | 0.3990130 | 0.417035 | 0.28 | 0.401171 | 0.418408 | -0.07 |
| rs671 | *ALDH2* | 12 | 111803962 | 0.0015270 | 0.000061 | -2.35 | 0.001300 | 0.000064 | -2.15 |
| rs2228093 | *ALDH1B1* | 9 | 38396005 | 0.1333550 | 0.128766 | -0.40 | 0.131952 | 0.129113 | -0.25 |
| rs2073478 | *ALDH1B1* | 9 | 38396068 | 0.4095980 | 0.405500 | 0.02 | 0.407919 | 0.402344 | -0.35 |
| rs148754219 | *GRM3* | 7 | 86644771 | 0.0074010 | 0.007396 | -0.03 | 0.007156 | 0.008067 | 0.78 |

For the previously implicated genes listed in the introduction and for *VGF, FOXP1, ARHGAP33* and *CDH9* additional analyses were carried out as described in order to see if there were particular categories of variant associated with the phenotypes. With a single exception, no category of variant within any of these genes showed evidence for association with heavy or problem drinking. The exception is that disruptive variants in *ADH1C* initially appeared to be protective for both heavy drinking, SLP = −3.02, OR = 0.73 (0.60–0.88), and for problem drinking, SLP = −3.79, OR = 0.68 (0.56–0.83). Although there were 7 different disruptive variants, 6 of them were extremely rare and the effect was driven by a single stop-gained variant with MAF=0.0099, 4:99347033:C>A. When the other variants within the ADH gene cluster were included as covariates in a multivariate analysis then this association disappeared and only the effect of rs1229984 itself was significant, suggesting that the overall result was due to LD between the stop variant and rs1229984 rather than any separate effect from impaired functioning of *ADH1C*. The results for the analyses of categories of variant are presented in Supplementary Table S3.

## Discussion

No genes attain conventional levels of statistical significance after correction for multiple testing. The two well-established variants in *ADH1B* and *ALDH2*, rs1229984 and rs671, demonstrate association when tested for individually but there is no evidence to implicate other rare coding variants in alcohol-metabolising genes. Overall, the conclusion is that this large sample of exome sequenced subjects has failed to identify additional genes or variants influencing susceptibility to heavy drinking or problem drinking.

Both phenotypes used clearly have their limitations. The heavy drinking phenotype is based on a report of drinking over the last 24 hours and so clearly might be more prone to random fluctuations than one based on a self-reported average consumption over a week or month, as was used in the previous study (Thompson et al., 2020). On the other hand, it might have an advantage in terms of being less sensitive to recall and reporting bias. Likewise, the phenotype for problem drinking depended on answering positively to any of a number of questions covering a wide range of severities from being addicted to alcohol to having had somebody suggest cutting down. Nevertheless, the fact that both phenotypes produced highly significant results with rs1229984 supports the thesis that they do have some validity. In fact, it is noteworthy that the effect size for this variant seems very similar for both phenotypes. This appears to be the case also for rs671 although its rarity in this dataset makes it difficult to draw firm conclusions.

Although no gene reached formal significance levels it is possibly of some interest that three genes potentially related to autism were each individually significant at p < 0.001: *FOXP1, ARHGAP33* and *CDH9*. There is also a report claiming to show association with alcoholism of clustered SNPs near other cadherin genes, *CDH11* and *CDH13*, though this was not genome-wide significant, and we have previously reported a statistically significant association between alcoholism in bipolar disorder and common variants in *CDH11* and (Johnson et al., 2006; Lydall et al., 2011). There is a negative association between alcohol abuse and autism and adults with autism spectrum disorders have relatively low rates of alcohol and substance use disorders (Vohra et al., 2017). Thus it would be easy to speculate that if there were genetic variants associated with autistic traits then these might be protective against heavy drinking and problem drinking. However a study of the polygenic risk for general substance abuse failed to detect a correlation with genetic susceptibility to autism, although this may be a feature of the relatively small sample size used (Carey et al., 2016). The other gene noted to be of potential interest, given its claimed involvement in psychiatric phenotypes, is *VGF*. However it needs to be stressed that none of these findings is formally significant and they should be regarded merely as hypothesis-generating.

Although the UK Biobank sample consists of 500,000 participants, to date exome sequence data has only been generally released for 200,000. It is possible that more definitive results may be obtained when the approach described here can be applied to the remaining participants and in other datasets. It is also possible that studying a more extreme phenotype such as alcohol dependence disorder would have more power but this would involve focussed recruitment strategies rather than relying on biobank resources. However at present it is not clear whether or not further genetic research will be able to yield actionable insights into factors influencing susceptibility to alcohol related disorders.

## Data Availability

The raw data is available on application to UK Biobank. Scripts and relevant derived variables will be deposited in UK Biobank. Software and scripts used to carry out the analyses are also available at https://github.com/davenomiddlenamecurtis.

## Acknowledgments

This research has been conducted using the UK Biobank Resource. The author wishes to acknowledge the staff supporting the High Performance Computing Cluster, Computer Science Department, University College London. This work was carried out in part using resources provided by BBSRC equipment grant BB/R01356X/1.

